# Sensitive quantitative detection of SARS-CoV-2 in clinical samples using digital warm-start CRISPR assay

**DOI:** 10.1101/2020.11.21.20236109

**Authors:** Xiong Ding, Kun Yin, Ziyue Li, Maroun M. Sfeir, Changchun Liu

## Abstract

Quantifying severe acute respiratory syndrome coronavirus 2 (SARS-CoV-2) in clinical samples is crucial for assessing the infectivity of coronavirus disease 2019 and the efficacy of antiviral drugs. Here, we describe a digital warm-start CRISPR (WS-CRISPR) assay for sensitive quantitative detection of SARS-CoV-2 in clinical samples. The WS-CRISPR assay combines low-temperature reverse transcription dual-priming mediated isothermal amplification (RT-DAMP) and CRISPR-Cas12a-based detection in one-pot, attributed to the mediation role by pyrophosphatase and phosphorothioated primers. The WS-CRISPR assay is initiated at above 50 °C and overcomes undesired premature target amplification at room temperature, enabling accurate digital nucleic acid quantification. By targeting SARS-CoV-2’s nucleoprotein gene, digital WS-CRISPR assay is able to detect down to 5 copies/μl SARS-CoV-2 RNA in the chip within 90 minutes. It is clinically validated by quantitatively determining 32 clinical swab samples and three clinical saliva samples, showing 100% agreement with RT-PCR results. Moreover, the digital WS-CRISPR assay has been demonstrated to directly detect SARS-CoV-2 in heat-treated saliva samples without RNA extraction, showing high tolerance to inhibitors. Thus, the digital WS-CRISPR method, as a sensitive and reliable CRISPR assay, facilitates accurate SARS-CoV-2 detection toward digitized quantification.

## Main

Since firstly emerged in December 2019^1^, severe acute respiratory syndrome coronavirus 2 (SARS-CoV-2) as the causing agent of coronavirus disease 2019 (COVID-19) has spread worldwide, resulting in over one million deaths^2^. As of now, fully validated COVID-19 vaccines and antiviral drugs are still unavailable. Therefore, sensitive and accurate quantification of the SARS-CoV-2 plays a crucial role in assessing the infectivity and controlling the ongoing pandemic.

To quantify SARS-CoV-2, TaqMan probe-based reverse transcription polymerase chain reaction (RT-PCR) is frequently used as a gold standard method due to its high sensitivity and specificity^3, 4^. However, it greatly depends on expensive real-time quantitative PCR instruments and its quantitation accuracy is highly associated with well-designed TaqMan probes, not suitable for small clinics or community health settings. Alternatively, some isothermal nucleic acid amplification methods have been developed to rapidly detect SARS-CoV-2, such as reverse transcription loop-mediated isothermal amplification (RT-LAMP)^5, 6, 7^, reverse transcription recombinase polymerase amplification (RT-RPA)^8, 9^, reverse transcription recombinase-aided amplification (RT-RAA)^10, 11^, and sensitive splint-based one-pot isothermal RNA detection (SENSR)^12^. However, most of these isothermal amplification assays are either intended for the qualitative detection or subjected to undesired nonspecific amplification signals (or false positive signals).

As next-generation molecular diagnostics, nucleic acid detection based on clustered regularly interspaced short palindromic repeats (CRISPR) and its associated proteins (Cas nucleases) possesses great prospects^13^. In CRISPR-Cas-based nucleic acid detection, target-specific CRISPR RNA (crRNA) ensures high specific and reliable detection and Cas nucleases’ collateral cleavage activity produces amplified fluorescence signals^14, 15, 16^. Currently, CRISPR-Cas12a-based DETECTR (DNA Endonuclease-Targeted CRISPR Trans Reporter) system and CRISPR-Cas13a-based SHERLOCK (Specific High-sensitivity Enzymatic Reporter UnLOCKing) system have been applied to detect SARS-CoV-2^17, 18^. However, these assays are typically two-step approaches in which RT-RPA or RT-LAMP, as separate target preampification step, is indispensable. The transferring of amplified products potentially increases the risk of carry-over contaminations and compilates the detection operation. To overcome this technical bottleneck, our lab and Zhang’s lab developed all-in-one dual CRISPR-Cas12a (AIOD-CRISPR) assay^19^ and SHERLOCK testing in one pot (STOP) assay^20^ for SARS-CoV-2 detection, respectively. However, these one-pot CRISPR assays are not quantitative. Recently, Yu’s lab has reported a digital quantitative strategy based on RPA-based CRISPR assay (termed RADICA) for synthetic SARS-CoV-2 RNA detection^21^. However, it is still a challenge to prevent undesired premature target amplification and accurately quantify nucleic acid, since the RPA/RT-RPA can be initiated at room temperature^22^, thereby potentially overestimating the initial target amount in the digital detection. To minimize premature target amplification, the reaction mixture needs to be prepared on ice and loaded into chips within one minute^21^, which complicates detection procedure and remains a challenge in massive COVID-19 testing.

Here, we present, for the first time, a digital warm-start CRISPR (WS-CRISPR) assay for sensitive quantitative detection of SARS-CoV-2 in clinical COVID-19 samples. The digital WS-CRISPR assay is established through partitioning a one-pot WS-CRISPR reaction into sub-nanoliter aliquots using QuantStudio 3D digital chips. The WS-CRISPR reaction combines low-temperature reverse transcription dual-priming isothermal amplification (RT-DAMP)^23^ and CRISPR-Cas12a-based fluorescence detection in a one-pot format and is efficiently initiated at above 50 °C, thereby preventing premature target amplifications at room temperature and enabling accurate digital quantification of nucleic acids. Through targeting the SARS-CoV-2’s nucleoprotein (N) gene, digital WS-CRISPR assay is developed to quantitatively determine 32 clinical swab samples and three clinical saliva samples. More importantly, digital WS-CRISPR assay is used to directly detect SARS-CoV-2 in heat-treated saliva samples without need for RNA extraction.

## Results

### Overview of digital WS-CRISPR assay

As shown in **Fig. 1a**, one-pot WS-CRISPR reaction mixture is first prepared in one tube, containing Cas12a-crRNA complex, six DAMP primers (two outer primers of FO and RO, two inner primers of FI and RI, and two competition primers of FC and RC), nontarget single-stranded DNA fluorophore-quencher (ssDNA-FQ) reporter, SuperScript IV reverse transcriptase, *Bst* DNA polymerase, and SARS-CoV-2 RNA target. To achieve digital detection, the prepared reaction mixture is distributed into a QuantStudio 3D digital chip (Thermo Fisher Scientific). This chip is etched with 20,000 consistently-sized hexagon wells (max width of 60 μm) in a silicon substrate and can partition the mixture into over ten thousand sub-nanoliter (∼0.7 nL) microreactions. After 90-min incubation at 52 °C, the microreactions with target RNA indicate strong green fluorescence (positive spots), whereas not in those without targets (negative spots). The target RNA can be quantified through testing and counting the number of positive spots of the chips.

**Fig. 1.**
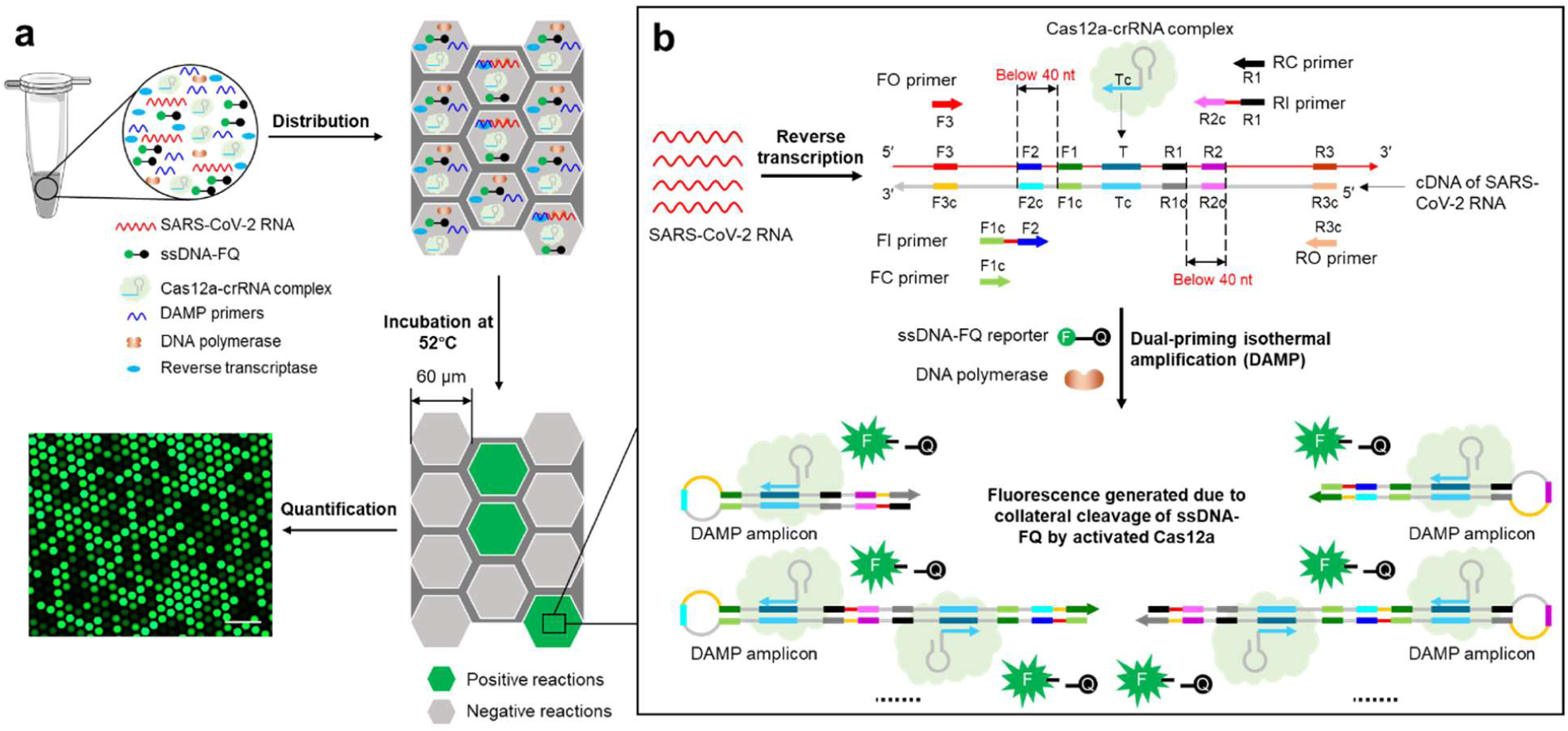
Overview of digital WS-CRISPR assay. **a**, One-pot WS-CRISPR reaction mixture is first prepared in one tube. After distributed into QuantStudio 3D digital chip, over ten thousand sub-nanoliter (∼0.7 nL) microreactions are isolated in microwells. When incubated at 52°C, each microreaction with SARS-CoV-2 RNA target undertakes WS-CRISPR reaction and generates strong green fluorescence (positive spots), whereas not in those without target (negative spots). Scale bar is 300 μm. Through detecting and counting the positive microreactions (or spots), SARS-CoV-2 RNA can be quantified based on the proportion of positive spots. **b**, Working principle of one-pot WS-CRISPR assay for SARS-CoV-2 detection. The WS-CRISPR reaction mixture contains Cas12a-crRNA complex, six DAMP primers (two outer primers of FO and RO, two inner primers of FI and RI, and two competition primers of FC and RC), ssDNA-FQ reporter, SuperScript IV reverse transcriptase, *Bst* DNA polymerase, and SARS-CoV-2 RNA in a one-pot format.

The WS-CRISPR assay combines low-temperature RT-DAMP with CRISPR-Cas12a detection in one-pot. The DAMP developed in our lab is one variant of loop-mediated isothermal amplification using a new primer design strategy^23^. Each inner primer of the DAMP is formed with two target sites with a distance of below 40 nt and a pair of competition primers is added to mediate pair-priming strand extension (**Fig. 1b**). Our previous study^23^ has confirmed that the DAMP/RT-DAMP has not only improved the detection sensitivity, but also generated ultralow nonspecific signals. In the digital WS-CRISPR assay, six DAMP primers recognize six distinct sites in target sequences to initiate self-priming and pair-priming (dual-priming) isothermal amplification, thus producing multiple amplicons with closed loop structures. Simultaneously, Cas12a-cRNA complex specifically binds the sites of the amplicons to activate Cas12a’s collateral cleavage activity, thereby indiscriminately cleaving surrounding nontarget ssDNA-FQ reporter to generate increased fluorescence. The ssDNA-FQ is a 5-cytosin nucleotide single-stranded DNA (5′-CCCCC-3′) labeled with FAM (Fluorescein) at 5′ end and Iowa Black FQ quencher at 3′ end, and is used in our assay due to its higher affinity to Cas12a^24^. Fluorescence is quenched via resonance energy transfer in intact ssDNA-FQ reporters, but can be recovered after activated Cas12a cleaves the reporters.

### Optimization of WS-CRISPR assay

As of now, there still remains a challenge to directly couple LAMP or DAMP with CRISPR-Cas12a detection in a one-pot format due to the significant difference in their reaction buffer compositions and reaction temperature. For example, one of the major concerns is the concentration of Mg^2+^. The cleavage of Cas12a nucleases for both on-target and collateral activity is typically high-Mg^2+^-dependent^25, 26, 27^. To enable highly sensitive nucleic acid detection, two different Cas12a nucleases were evaluated and compared at various Mg^2+^ concentrations. As shown **Fig. 2a**, when reducing the Mg^2+^ concentration from 8 mM to 2 mM, the detection efficiency of the CRISPR-Cas12a dramatically decreases. Interestingly, the Cas12a from *Recombinant Acidaminococcus sp. BV3L6* (*A*.*s*. Cas12a) still has relatively high collateral cleavage activity at 2 mM Mg^2+^. Whereas, 2 mM Mg^2+^ completely inhibits the activity of Cas12a from *Lachnospiraceae bacterium ND2006* (*Lba* Cas12a). Thus, *A*.*s*. Cas12a is used in our WS-CRISPR assay due to its tolerance to low Mg^2+^ concentration. In addition, during isothermal amplification, primer extension by DNA polymerase continuously consumes dNTPs and produces a large number of pyrophosphate ions that can chelate Mg^2+^ to form magnesium pyrophosphate precipitate as the reaction byproduct (**Fig. 2b**), thereby consuming a large amount of free Mg^2+^ and significantly reducing the collateral cleavage activity of CRISPR-Cas12a nuclease. To this end, pyrophosphatase (PPase) is added into the WS-CRISPR reaction system to degrade the magnesium pyrophosphate precipitate, which releases free Mg^2+^ and maintains a constant Mg^2+^ concentration. As shown in **Supplementary Fig. 1**, the optimal concentration of PPase is 0.2 U/μl in our WS-CRISPR assay.

**Fig. 2.**
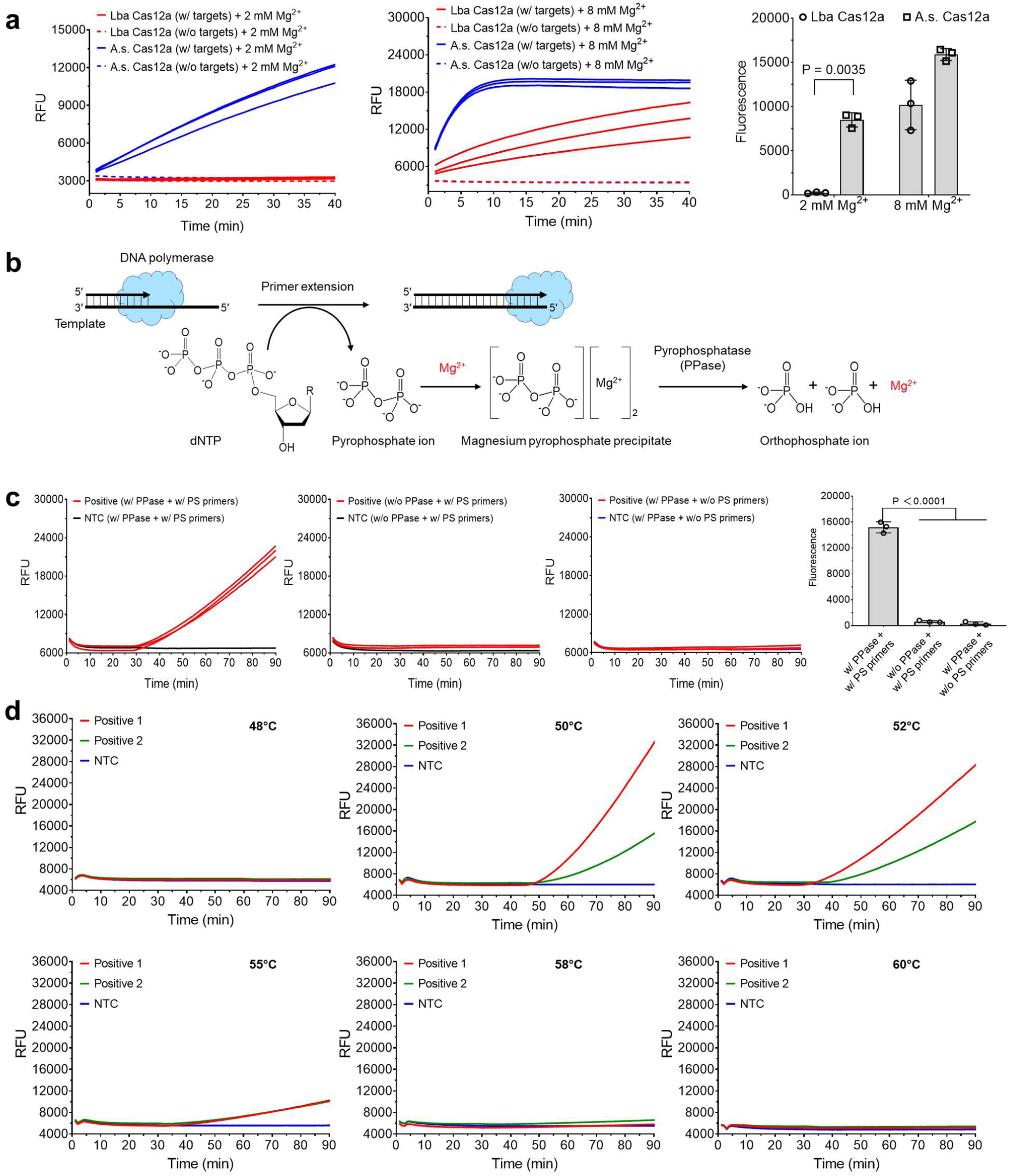
Optimization of one-pot WS-CRISPR assay. **a**, Comparison of two different Cas12a nucleases in one-pot WS-CRISPR assay at different Mg^2+^ concentration. Lba Cas12a, EnGen Lba Cas12a from *Lachnospiraceae bacterium ND2006* (New England Biolabs). A.s. Cas12a, Alt-R Cas12a (Cpf1) *Ultra* nuclease from *Recombinant Acidaminococcus sp. BV3L6* (Integrated DNA Technologies). The used targets are 1 μM of synthetic SARS-CoV-2 N DNA fragments. Three replicates were run (n = 3). **b**, The chemical reaction process of the generation and degradation of the magnesium pyrophosphate precipitate due to the existence of pyrophosphatase during primer extension or DNA polymerization. **c**, Real-time fluorescence detection and endpoint fluorescence comparison of one-pot WS-CRISPR assay with PPase and/or PS primers at 52°C. PS primers specifically denote two phosphorothioated inner primers of FI and RI and the rest of primers are non-phosphorothioated. “w/o PS primers” means reactions with the non-phosphorothioated inner primers. Positive, the reaction with 5×10^4^ copies/μl SARS-CoV-2 RNA. Three replicates were run (n = 3). **d**, Effect of reaction temperature on one-pot WS-CRISPR assay. Positive 1 and 2, the reactions with 3×10^6^ and 5×10^4^ copies/μl SARS-CoV-2 RNA, respectively. Three independent assays were conducted with the similar results. NTC, non-template control. Error bars represent the means ± standard deviation (s.d.) from replicates. The statistical significance was analyzed using unpaired two-tailed *t*-test.

Another concern in developing one-pot CRISPR assay is the significant difference of the optimal reaction temperature between nucleic acid amplification and CRISPR-based detection. Most Cas12a nucleases have an optimal activity at 37°C, but LAMP/DAMP powered by *Bst* DNA polymerase typically requires a relatively high temperature of 60-65°C^23, 28^. To develop a one-pot assay, phosphorothioated inner primers of FI and RI (**Supplementary Table 1**) were employed in our WS-CRISPR assay to reduce the reaction temperature of isothermal amplification as reported by Ellington’s lab^29^. Therefore, through supplementing PPase and employing phosphorothioated inner primers, one-pot WS-CRISPR assay has been successfully developed (**Fig. 2c**).

To achieve the best performance, we further optimize the one-pot WS-CRISPR reaction system in terms of the concentrations of Mg^2+^, *Bst* DNA polymerase, and SuperScript IV reverse transcriptase. As shown in **Supplementary Figs. 2-4**, the optimal concentrations are 0.2 U/μl PPase, 2 mM Mg^2+^, 24 U/μl *Bst* DNA polymerase, and 2 U/μl SuperScript IV. In addition, several different DNA polymerases were investigated and compared, including *Bst* DNA polymerase (large fragment), *Bst* 2.0 DNA polymerase, *Bst* 3.0 DNA polymerase, GspSSD 2.0 DNA polymerase, *Bsm* DNA polymerase (large fragment), IsoPol BST^+^ DNA polymerase, and IsoPol SD^+^ DNA polymerase. As shown in **Supplementary Fig. 5**, the best DNA polymerase for our WS-CRISPR is *Bst* DNA polymerase (large fragment). Furthermore, we assessed the effect of reaction temperatures from 48 °C to 60 °C on the one-pot WS-CRISPR assay. **Fig. 2d** shows that the optimal temperature of the WS-CRISPR assay is 52 °C and the reaction is initiated at above 50 °C. Therefore, our WS-CRISPR assay provides a warm-start nucleic acid detection and circumvents undesired premature target amplification at room temperature, which is crucial to developing a sensitive and reliable digital detection for nucleic acid quantification.

As shown in **Fig. 3a**, the WS-CRISPR reaction is only initiated when all the components of the RT-DAMP and CRISPR-Cas12a are mixed in one-pot. In addition, the specificity of WS-CRISPR assay is evaluated by detecting SARS-CoV control, MERS-CoV, and Hs_RPP30 control. As shown in **Fig. 3b**, the WS-CRISPR assay has a high specificity to detect SARS-CoV-2. By detecting various concentrations of SARS-CoV-2 RNA, the WS-CRISPR assay is able to detection down to 500 copies/μl SARS-CoV-2 RNA in sample (equivalently 50 copies/μl RNA in the reaction) within 90 min (**Fig. 3c**). Furthermore, **Fig. 3** demonstrates that the detection results of the WS-CRISPR assay can be visually read out based on the fluorescence imaging of reaction tubes under either LED blue light or UV light, enabling simple visual detection.

**Fig. 3.**
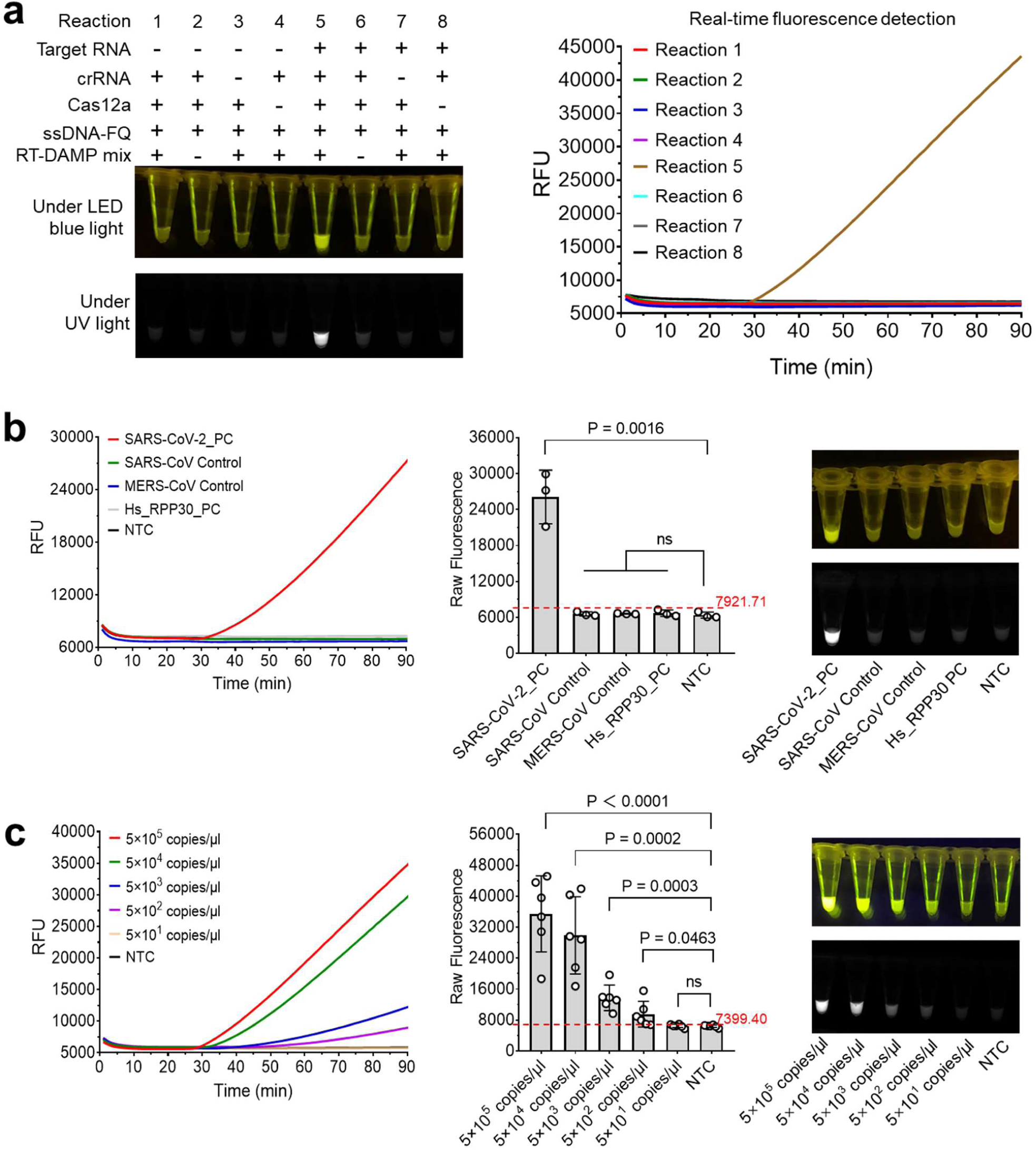
Specificity and sensitivity of one-pot WS-CRISPR assay for SARS-CoV-2 detection. **a**, Evaluation of eight reactions with various components through endpoint imaging after 90-min incubation and real-time fluorescence detection. Target RNA, 5×10^5^ copies/μl SARS-CoV-2 RNA. RT-DAMP mix contains six primers, reverse transcriptase, and *Bst* DNA polymerase in one-pot reaction. **b**, Specificity of real-time/endpoint one-pot WS-CRISPR assay for SARS-CoV-2 detection. Three replicates were run (n = 3). **c**, Sensitivity of real-time/endpoint WS-CRISPR assay when detecting various concentrations of SARS-CoV-2 RNA. Six replicates were run (n = 6). NTC, non-template control. Horizontal dashed lines indicate the cutoff fluorescence defined as the average intensity of NTC plus three times of the standard deviation. Error bars represent the means ± s.d. from replicates. The statistical significance was analyzed using unpaired two-tailed *t*-test.

In summary, we successfully develop a one-pot warm-start CRISPR assay by combining the low-temperature RT-DAMP and CRISPR-Cas12a-based fluorescence detection in a one-pot format. It has high sensitivity and specificity for SARS-CoV-2 detection in both real-time fluorescence monitoring and endpoint visual readout. Especially, the WS-CRISPR assay is typically initiated at above 50°C, presenting the first report of one-pot warm-start CRISPR-Cas12a assay, which is crucial to develop a digital CRISPR assay for accurate and reliable quantification of nucleic acids.

### Development of digital WS-CRISPR assay

The digital WS-CRISPR assay is developed through partitioning the one-pot WS-CRISPR reaction mixture into sub-nanoliter microreactions in the QuantStudio 3D digital chips. As shown in **Fig. 4a**, a typical workflow of digital WS-CRISPR assay consists of RNA extraction from clinical samples, one-pot CRISPR reaction mixture preparation, distribution of the reaction mixture into the chip, and incubation at 52°C. First, the digital WS-CRISPR assays with various incubation times (e.g., 10, 30, 60, 90 and 120 min) were investigated. As shown in **Fig. 4b** and **4c**, a 90-min incubation is enough for the digital WS-CRISPR assay to reach the maximum percentage of positive spots.

**Fig. 4.**
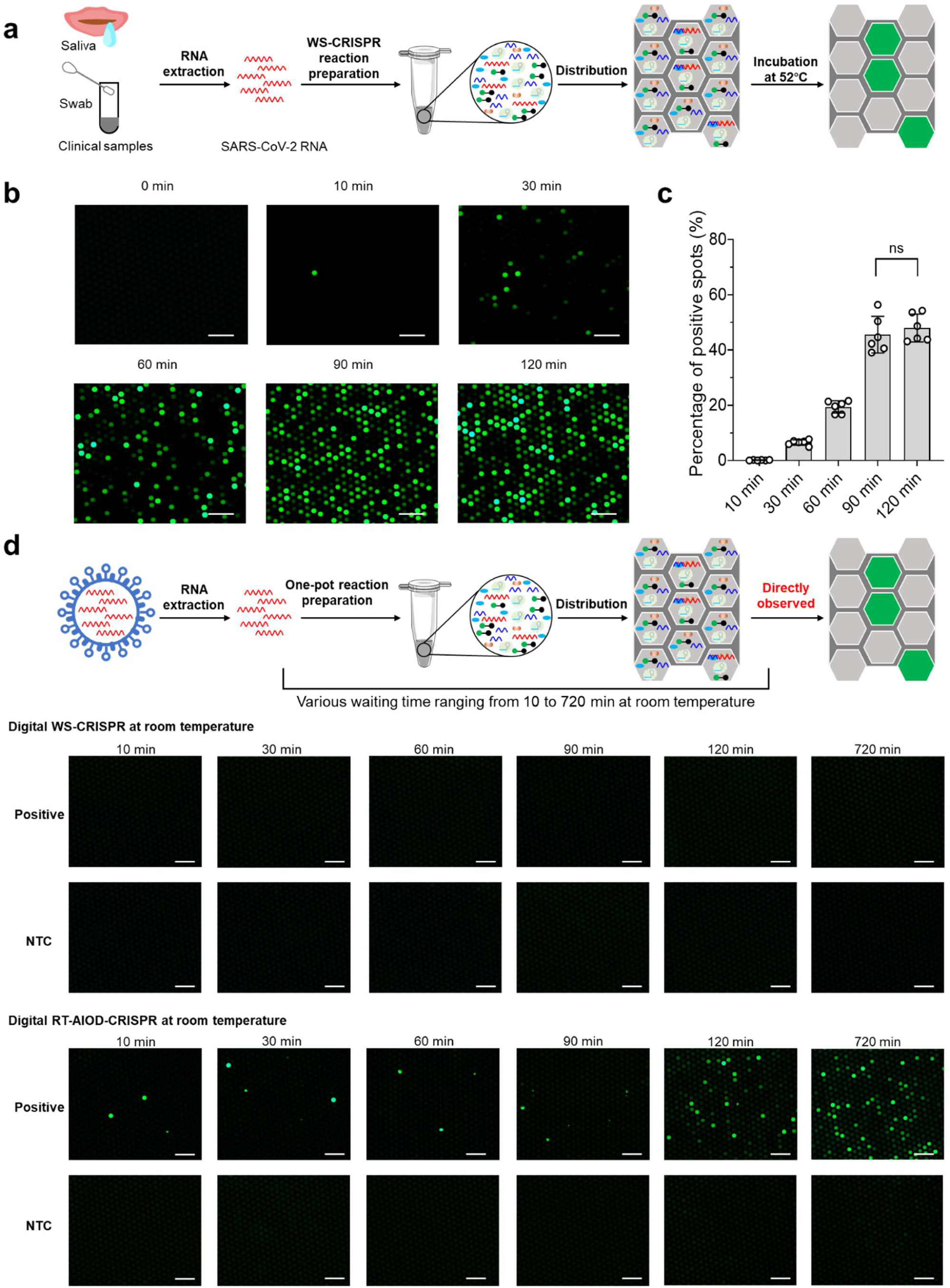
Digital WS-CRISPR assay for SARS-CoV-2 detection. **a**, A typical workflow of digital WS-CRISPR assay to detect SARS-CoV-2 in clinical samples. **b**, Endpoint fluorescence micrographs of the QuantStudio digital chip for the SARS-CoV-2 detection with various incubation time (0, 10, 30, 60, 90 and 120 min) at 52°C. In this digital WS-CRISPR assay, 1×10^6^ copies/μl SARS-CoV-2 RNA was loaded. **c**, The percentage of positive spots comparison for the digital WS-CRISPR assays with various incubation time at 52°C.The number of positive spots was counted by setting the same threshold in the ImageJ software. Percentages of positive spots in each micrograph was calculated (n = 6). Error bars represent the means ± s.d. from replicates. The statistical significance was analyzed using unpaired two-tailed *t*-test. **d**, Effect of various waiting time at room temperature on digital WS-CRISPR assay and digital RT-AIOD-CRISPR assay during reaction solution preparation and distribution steps. After specific waiting time at room temperature, the chips were directly observed without incubation. Positive, the reaction with 5×10^5^ copies/μl SARS-CoV-2 RNA. NTC, non-template control. Scale bars are 300 μm. Each micrograph is a representative of six distinct regions taken to cover about 2809 microreactions.

To avoid overestimating the initial amount of target nucleic acid by digital detection, it is crucial to prevent undesired premature target amplification during reagent preparation at room temperature. For example, some DNA polymerization reactions (e.g., RPA) can be initiated at room temperature^22^. To determine if there is any premature target amplification in our digital WS-CRISPR assay, we set up various waiting times at room temperature during the reaction solution preparation and distribution steps (**Fig. 4d**). For comparison, we also assess the digital detection of our previously reported RT-AIOD-CRISPR assay, a one-pot RT-RPA-based CRISPR assay (*19*). As shown in **Fig. 4d**, positive spots can clearly be observed as short as 10 min-waiting time at room temperature in the digital RT-AIOD-CRISPR assay, confirming the premature target amplification occurs, which is consistent to recent report with the RADICA assay in digital chips by Yu’s lab (*21*). To minimize premature target amplification, they prepared reaction mixture on ice and quickly distributed it into the chips (within 1 min)^21^. However, this complicates the assay’s procedure and typically requires highly well-trained operators. On the contrary, no positive spots are observed in our digital WS-CRISPR assay even after 720-min waiting time at room temperature (**Fig. 4d**). Thus, our digital WS-CRISPR method provides a warm-start assay strategy and enables a simple, sensitive, and reliable quantification of SARS-CoV-2.

Next, specificity assay of the digital WS-CRISPR is carried out by testing non-SARS-CoV-2 nucleic acids. As shown in **Fig. 5a**, positive spots are observed in the chip loaded with the SARS-CoV-2 positive control, whereas not for other non-target nucleic acids, such as SARS-CoV control, MERS-CoV, and Hs_RPP30 control, which is consistent to the results of the tube-based bulk reaction format (**Fig. 3b**). By testing various concentrations of SARS-CoV-2 RNA, the detection sensitivity is investigated. As shown in **Fig. 5b**, digital WS-CRISPR assay is able to detect down to 50 copies/μl SARS-CoV-2 RNA in the sample (equivalently 5 copies/μl RNA molecules in the reaction), a 10-fold higher compared with the bulk WS-CRISPR assay in the tube format (**Fig. 3c**). In addition, **Fig. 5b** also indicates that there is an excellent linear relationship (R^2^=0.9934) between the concentration of targets (from 5×10^3^ to 3×10^6^ copies/μl) and the percentage of positive spots. However, when plotting the number of positive spots in digital WS-CRISPR assays according to the commonly used Poisson distribution^30^, we find that the target concentration quantified by our digital WS-CRISPR assay is lower than that of the initial target added, likely attributed to relatively low filling rate of microwells in the digital chip. In our digital WS-CRISPR assay, we used commercially available QuantStudio digital chip that is specially designed for digital PCR/RT-PCR assay. The WS-CRISPR reaction system has a distinct wettability behavior on the digital chip due to the different reaction components. We will explore other digital chips (e.g., Clarity™ digital chip^21^) to test our digital WS-CRISPR assay in the future. Nonetheless, our digital WS-CRISPR has the capability of accurately quantifying targets based on a standard curve.

**Fig. 5.**
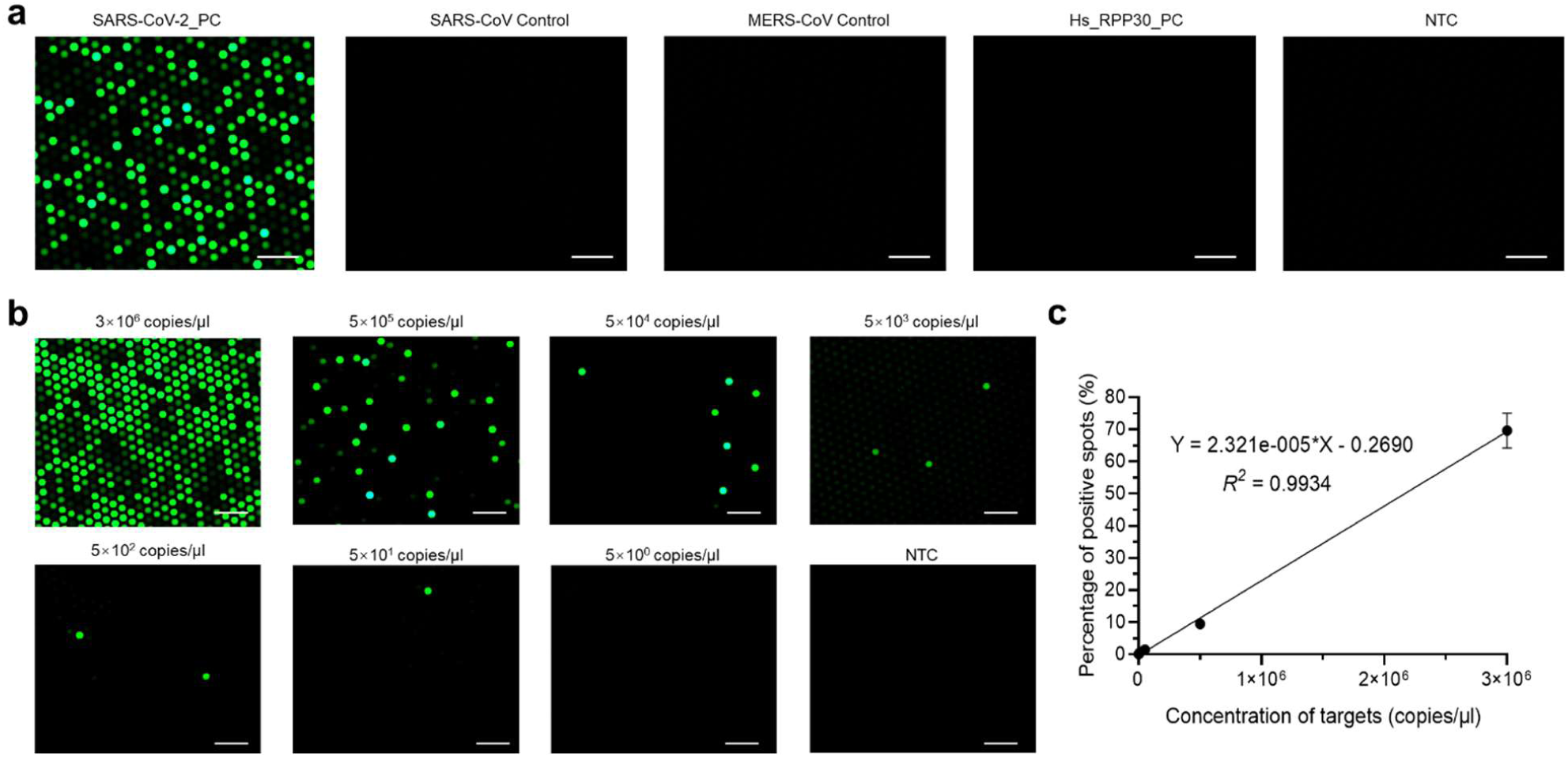
Specificity and sensitivity of digital WS-CRISPR assay for SARS-CoV-2 detection. **a**, Endpoint fluorescence micrographs of the QuantStudio digital chip for the specificity detection of the digital WS-CRISPR assays. SARS-CoV-2 PC, SARS-CoV control, MERS-CoV control, and Hs_RPP30 PC were from Integrated DNA Technologies. **b**, Endpoint fluorescence micrographs of the chip for the digital WS-CRISPR assays testing various concentrations of SRS-CoV-2 RNA within 90-min incubation at 52°C. **c**, The linear relationship between percentage of positive spots and concentration of targets. The number of positive spots was counted by setting the same threshold in the ImageJ software. For each concentration’s testing, total positive spots in all the six micrographs were used and three chips were taken to run three independent assays (n = 3). Each micrograph is a representative of six distinct regions taken to cover about 2809 microreactions. Scale bars are 300 μm. Error bars represent the means ± s.d. from replicates.

### Clinical validation of digital WS-CRISPR assay

To validate the clinical utility of the digital WS-CRISPR assay, we detected SARS-CoV-2 RNA extracted from 32 clinical swab samples and three clinical saliva samples. For comparison, an in-home RT-qPCR assay using the U.S. CDC-approved SARS-CoV-2 N1 gene’s primers and probes (provided by Integrated DNA Technologies) were set up as the parallel experiment. As shown in **Fig. 6a**, 12 positive samples and positive control are consistently detected and identified by digital WS-CRISPR assays, while all negative samples show negative signals, showing a 100% agreement with that of RT-qPCR method. **Fig. 6b** shows the determined concentrations of SARS-CoV-2 RNA in the 12 positive samples and the positive control by digital WS-CRISPR and RT-qPCR methods. The averaged viral loading was quantified with the range from 1.4×10^4^ to 2.3×10^6^ copies/μl, showing similar order of magnitude as those determined by RT-qPCR. However, **Fig. 6b** also indicates that digital WS-CRISPR cannot quantify the sample 12 and 19 with larger Cq values (Cq = 26.09 and 35.34) in RT-qPCR assay, likely attributed to its high limitation of quantitation (**Fig. 5c**). Despite it, the digital WS-CRISPR assay is able to quantify the SARS-CoV-2 RNA extracted from both clinical swab and saliva samples, showing a comparable performance with conventional RT-qPCR method. In addition, our digital WS-CRISPR method is the first digital CRISPR assay that is clinically validated by using clinical samples.

**Fig. 6.**
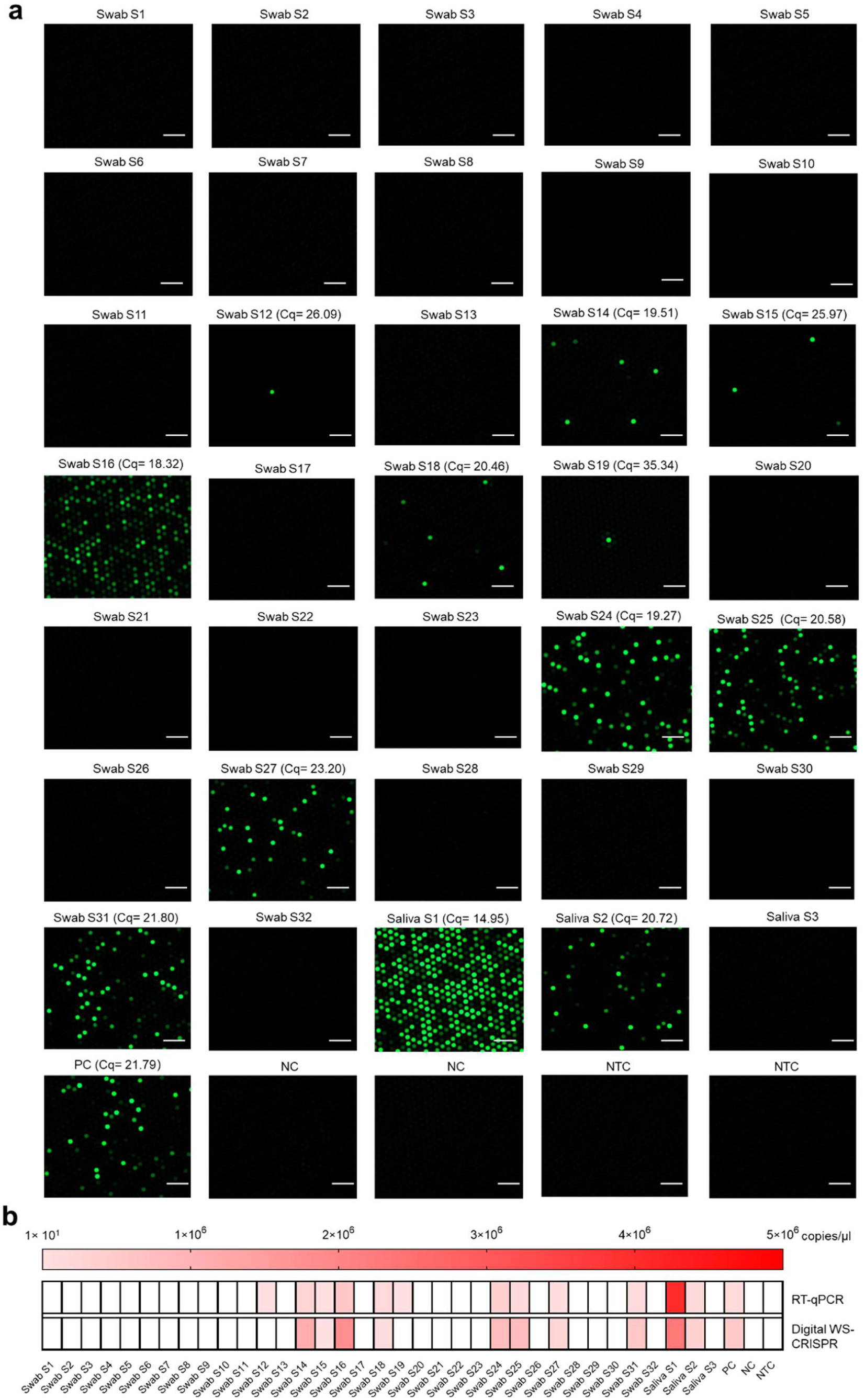
Clinical validation of digital WS-CRISPR assay. **a**, Endpoint fluorescence micrographs of the QuantStudio digital chip for detecting SARS-CoV-2 RNA extracted from 32 clinical swab samples (Swab S1-S32) and three saliva samples (Saliva S1-S3). The indicated Cq values were the results of RT-qPCR assays. **b**, Heat map displaying the determined SARS-CoV-2 RNA concentration by RT-qPCR and digital WS-CRISPR for each sample. The presented concentrations are the average values in three independent assays. Blank means SARS-CoV-2 negative. Each micrograph is a representative of six distinct regions taken to cover about 2809 microreactions. PC, SARS-CoV-2-positive control sample. NC, SARS-CoV-2-negative control sample. NTC, non-template control. Scale bars are 300 μm.

### Direct SARS-CoV-2 detection in saliva samples by digital WS-CRISPR assay

Recent research showed that saliva sampling is an attractive alternative to swab sampling in SARS-CoV-2 detection due to its simplicity, convenience and non-invasive nature^31^. Especially, saliva samples can be self-collected by patients themselves, avoiding direct interaction between health care workers and patients. Given this, we investigated whether our digital WS-CRISPR assay can directly be adapted to detect SARS-CoV-2 in crude saliva samples without RNA extraction step. As shown in **Fig. 7a**, each saliva sample contains 90% (v/v) of human saliva obtained from healthy individual, 0%-10% (v/v) of spiked heat-inactivated SARS-CoV-2 from BEI Resources (Catalog # NR-52350), and 1× inactivation reagent developed by Rabe and Cepko^7^. After heated at 95°C for 5 min, 1.5 μl of the saliva samples was directly added into the digital WS-CRISPR reaction system. As shown in **Fig. 7b**, the digital WS-CRISPR assay is successfully able to detect the SARS-CoV-2 spiked in the saliva samples without need for RNA extraction and purification, exhibiting high tolerance to potential inhibitors in saliva samples due to reaction partitioning. Thus, this interesting finding suggests that our digital WS-CRISPR assay has the potential to directly detect SARS-CoV-2 from crude clinical saliva specimens through simple heating treatment, facilitating rapid and early molecular diagnostics of COVID-19 infection.

**Fig. 7.**
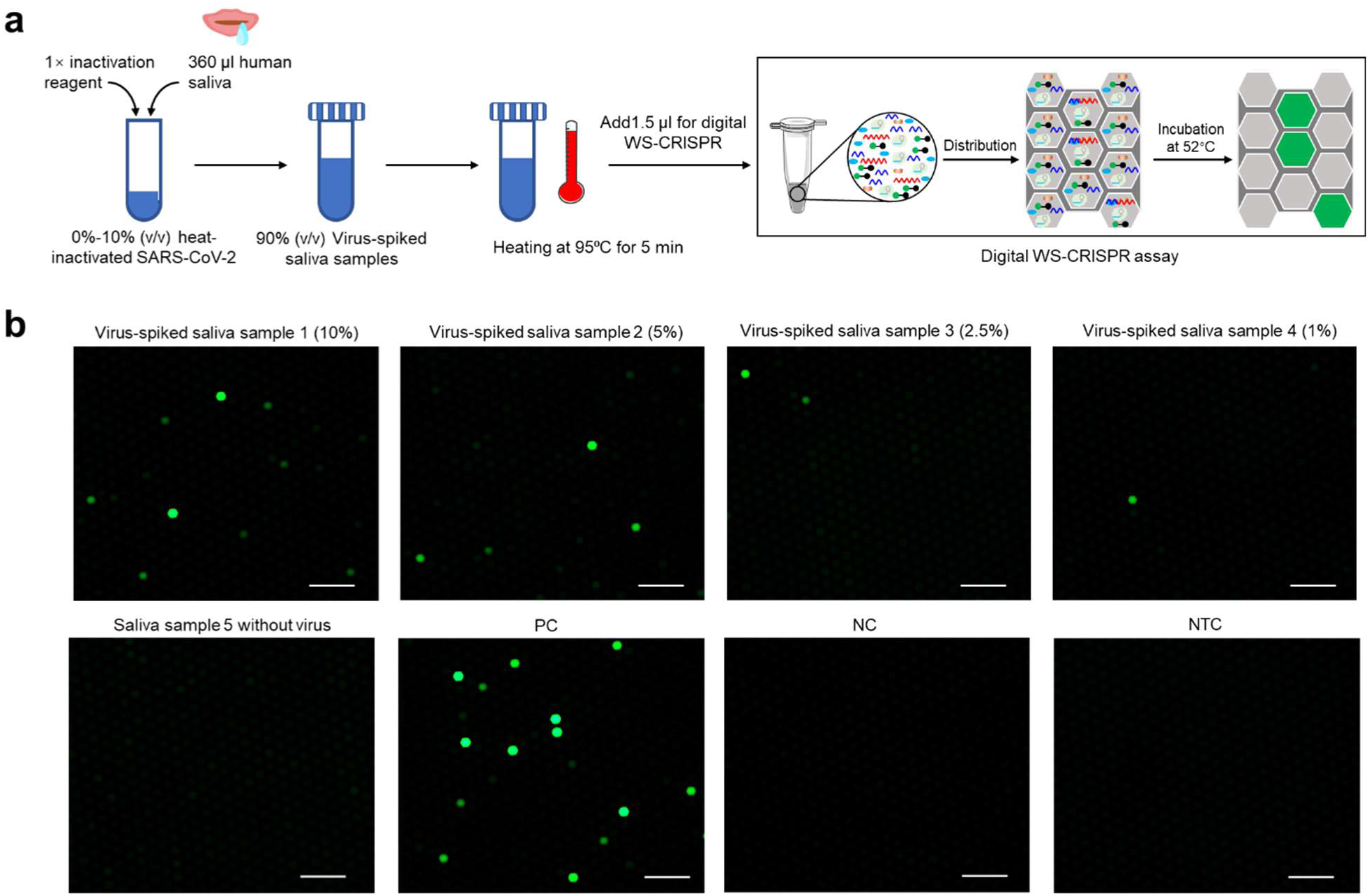
Direct detection of SARS-CoV-2 in crude saliva samples by digital WS-CRISPR assay. **a**, Workflow for direct SARS-CoV-2 testing in spiked saliva samples by digital WS-CRISPR assay. **b**, Endpoint fluorescence micrographs of the chip for direct detection of SARS-CoV-2 virus spiked in saliva samples. Saliva samples 1-5, the samples with 10%, 5%, 2.5%, 1%, and 0% of heat-inactivated SARS-CoV-2 virus. Each micrograph is a representative of six distinct regions taken to cover about 2809 microreactions. PC, SARS-CoV-2-positive control sample. NC, SARS-CoV-2-negative control sample. NTC, non-template control. Scale bars are 300 μm.

## Discussion

In this study, we proposed a digital WS-CRISPR-Cas12a assay for sensitive quantitative detection of SARS-CoV-2 from clinical samples. We took advantage of a newly established one-pot warm-start CRISPR reaction combining a low-temperature RT-DAMP and CRISPR-Cas12a-based detection. To couple these two different reaction systems into one-pot, pyrophosphatase is added to maintain a constant Mg^2+^ concentration by degrading the magnesium pyrophosphate byproduct. In addition, phosphorothioated inner primers are used to mediate efficient reverse transcription isothermal amplification at relatively low-temperature such as 52°C. Through partitioning the one-pot reaction mixture into sub-nanoliter microreactions using QuantStudio 3D digital chips, we successfully develop a digital CRISPR assay for sensitive and reliable quantification of SARS-CoV-2.

Compared to previously reported CRISPR-based nucleic acid assays^17, 18, 19, 20, 21^, our digital WS-CRISPR assay offers several remarkable advantages. First, our WS-CRISPR assay is the first demonstration of one-pot CRISPR assay by combining *Bst* DNA polymerase-based reverse transcription isothermal amplification with CRISPR-Cas12a detection, not relying on CRISPR-Cas12b that requres higher reaction temperature (e.g., 60-65 °C) and longer crRNA (> 100 nt)^32, 33^. Second, digital WS-CRISPR assay is typically initiated at an elevated temperature (e.g., above 50°C), thoroughly addressing the challenge of undesired premature target amplification in digital detection. Third, digital WS-CRISPR assay has high detection specificity and 10-fold higher sensitivity than tube-based bulk assay format. By targeting the SARS-CoV-2’s N gene, digital WS-CRISPR assay is able to quantify down to 5 copies/μl SARS-CoV-2 RNA in the chip. Fourth, digital WS-CRISPR assay can quantify the SARS-CoV-2 in clinical samples, benefiting assessing the COVID-19 infectivity and the efficacy of antiviral drugs. Last, our digital WS-CRISPR assay shows high tolerance to inhibitors and can directly detect SARS-CoV-2 in crude saliva samples without need for RNA extraction, which does not only facilitate the COVID-19 diagnosis, but also lowers the infection risk in health workers without directly sampling from patients.

These advantages notwithstanding, new digital chip needs to be further explored for digital WS-CRISPR assay in the future, enabling absolute quantitative of nucleic acids. In addition, although our digital WS-CRISPR assay uses relatively expensive fluorescence microscopy for fluorescence image, smartphone-based portable fluorescence microscopy can become the alternative to achieve onsite quantitative detection^34, 35, 36^. Of note, digital WS-CRISPR opens a new exploration for CRISPR-base nucleic acid quantitative detection. For example, although RT-DAMP is only used in this study, other isothermal methods are also applicable with our proposed strategies, such as RT-LAMP, reverse transcription isothermal multiple-self-matching-initiated amplification (RT-IMSA)^37^, and reverse transcription cross-priming amplification (RT-CPA)^38^. As the first clinically validated digital CRISPR assay, our digital WS-CRISPR assay provides a reliable, sensitive and straightforward SARS-CoV-2 quantitative detection.

## Methods

### Reagents and samples

Bovine serum albumin (BSA, 20 mg/ml), EnGen Lba Cas12a (100 µM), deoxynucleotide (dNTP) mix (10 mM of each), RNase inhibitor (Murine, 40,000 U/ml), extreme thermostable single-stranded DNA binding protein (ET-SSB, 500 µg/ml), isothermal amplification buffer pack (10×; containing 200 mM Tris-HCl, 500 mM KCl, 100 mM (NH4)2SO4, 20 mM MgSO4, 1% Tween 20, and pH = 8.8 at 25°C), thermostable inorganic pyrophosphatase (PPase, 2,000 U/ml), *Bst* DNA polymerase (large fragment), *Bst* 2.0 DNA polymerase, *Bst* 3.0 DNA polymerase, and nuclease-free water were purchased from New England BioLabs (Ipswich, MA). GspSSD 2.0 DNA polymerase was from OptiGene (West Sussex, UK). IsoPol BST^+^ and IsoPol SD^+^ polymerases were purchased from ArcticZymes Technologies (Norway). Invertase from *Saccharomyces cerevisiae* (Grade VII, ≥300 U/mg), taurine (≥99%, 10 g), TCEP-HCl (Reagent Grade, 5 g), and NaOH (≥98%, 500 g) were purchased from Sigma-Aldrich (St. Louis, MO). UltraPure EDTA (0.5 M, pH = 8), *Bsm* DNA Polymerase, large fragment (8 U/µl), SuperScript IV reverse transcriptase (200 U/µL), QuantStudio 3D digital PCR 20K chip kit (Version 2), and digital PCR master mix were purchased from Thermo Fisher Scientific (Waltham, MA). Normal saliva human fluid was purchased from MyBioSource (San Diego, CA). Heat-inactivated SARS-CoV-2 (Isolate USA-WA1/2020, NR-52350) was from BEI Resources (Manassas, VA). Synthetic SARS-CoV-2 RNA control (MN908947.3) with a coverage of greater than 99.9% of the bases of the SARS-CoV-2 viral genome was purchased from Twist Bioscience (San Francisco, CA). Alt-R A.s. Cas12a *Ultra* (500 µg; 64 µM), SARS-CoV-2 positive control (SARS-CoV-2 _PC, Catalog # 10006625), SARS-CoV control (Catalog # 10006624), and Middle East respiratory syndrome coronavirus control (MERS-CoV control, Catalog # 10006623), human RPP30 gene control (Hs_RPP30_PC, Catalog # 10006626), nCOV_N1 Forward Primer Aliquot (50 nmol, Catalog # 10006821), nCOV_N1 Reverse Primer Aliquot (50 nmol, Catalog # 10006822), and nCOV_N1 Probe Aliquot (50 nmol, Catalog # 10006823) were purchased from Integrated DNA Technologies (Coralville, IA). 32 clinical swab samples and three clinical saliva samples were tested and their viral RNAs were extracted by utilizing QIAamp DSP Viral RNA Mini Kit (QIAGEN N.V., Venlo, The Netherlands). All clinical samples were de-identified and handled in compliance with ethical regulations and the approval of Institutional Review Board of the University of Connecticut Health Center (protocol #: P61067).

### Design of primers and crRNA

Six DAMP primers and one CRISPR-Cas12a’s crRNA were designed to target seven distinct sites in the 173 bp SARS-CoV-2 N gene fragment with the location from 28769 to 28941 in the viral genome (GenBank accession MW202218.1). The selected primer or crRNA recognition sites are highly conserved based on the GISAID-provided genomic epidemiology of hCoV-19 for 3564 genomes sampled between Dec 2019 and Nov 2020 (as of Nov 11, 2020. https://www.gisaid.org/epiflu-applications/phylodynamics/). The DAMP primers can be manually designed using the OligoAnalyzer Tool (https://www.idtdna.com/pages) and the PrimerExplorer (https://primerexplorer.jp/e/) according to previously reported design principle^23^. Also, they can be designed using the online DAMP primer design platform developed by our lab (https://github.com/xuzhiheng001/DAMP-Design). CRISPR-Cas12a’s crRNA targeting the middle site of DAMP region (**Fig. 1**) was also designed using the OligoAnalyzer Tool and its sequence was checked against MERS-CoV and SARS-CoV gene sequences. Primers, crRNA, ssDNA-FQ reporter, and SARS-CoV-2 N DNA fragments were synthesized from Integrated DNA Technologies (Coralville, IA). All the sequence information of the used primers and crRNAs has been listed in **Supplementary Table 1**.

### One-pot WS-CRISPR assay

One-pot WS-CRISPR assay system was prepared separately as Component A and B. Component A consisted of 1× isothermal amplification buffer (20 mM Tris-HCl, 50 mM KCl, 10 mM (NH4)2SO4, 2 mM MgSO4, 0.1% (v/v) Tween 20, and pH = 8.8 at 25°C), 2 U/μl SuperScript IV reverse transcriptase, 50 mM taurine, 1 U/μl invertase, 0.01 mg/ml BSA, 0.4 mM each of dNTPs, 0.2 μM FO primer, 0.2 μM RO primer, 1.6 μM PS-FI primer, 1.6 μM PS-RI primer, 1.6 μM FC primer, 1.6 μM RC primer, 10 μM ssDNA-FQ, 20 ng/μl ET SSB, 24 U/μl Bst DNA polymerase (large fragment), 0.2 U/μl PPase, and 1 U/μl RNase inhibitor. Component B contained 1.28 μM A.s. Cas12a (*Ultra*) and 1.2 μM DAMP-crRNA (**Supplementary Table 1**). All the indicated concentrations were calculated based on the finally assembled reaction system. In a typical 15-μl assay, 12.5 μl Component A was first mixed with 1.5 μl of the target solution and then supplemented with 1.0 μl Component B. The assembled reaction mixture was then incubated at 52°C for 90 min in the Bio-Rad CFX96 Touch Real-Time PCR Detection System (Hercules, CA) for real-time fluorescence detection. After incubation, the tubes were placed in the Maestrogen UltraSlim LED blue light illuminator (Pittsburgh, PA) or the Bio-Rad ChemiDoc MP Imaging System with its built-in UV channel (Hercules, CA) for endpoint visual detection. The endpoint fluorescence was the determined raw fluorescence subtracting the averaged raw fluorescence of non-template control reaction. Specificity assay was investigated by using the control plasmids containing the complete N gene from SARS-CoV-2_PC, SARS-CoV control, MERS-CoV control, and Hs_RPP30_PC. Sensitivity assay was conducted by testing serially diluted SARS-CoV-2 RNA (Twist Bioscience) in water with concentrations of 5×10^5^, 5×10^4^, 5×10^3^, 5×10^2^, and 5×10^1^ copies/μl. CRISPR-Cas12a-based detection, namely trans-cleavage assay was incubated at 37°C for 40 min and performed in a solution containing 1× isothermal amplification buffer, 0.32 μM A.s. Cas12a, 0.32 μM DAMP-crRNA, 1.0 μM synthetic SARS-CoV-2 N DNA fragment, and 1.0 μM ssDNA-FQ reporter.

### Digital WS-CRISPR assay

The reaction system for digital WS-CRISPR assay was the same as that in the tube-based bulk reaction mentioned above. The procedure of digital WS-CRISPR assay was modified based on the operational workflow for QuantStudio 3D digital PCR (Quick Reference Manual. Thermo Fisher Scientific). Briefly, a 15-μl digital WS-CRISPR reaction solution was first prepared in a tube by mixing Component A, B and the sample. Then, the reaction solution was loaded into the QuantStudio 3D digital PCR chip (version 2), followed by applying the lid, loading the immersion fluid, and sealing the chip. This step can be finished by using the QuantStudio 3D digital PCR Chip Loader (Thermo Fisher Scientific). Afterwards, the sealed chip was placed in ProFlex 2× Flat PCR System (Thermo Fisher Scientific) for 90-min incubation at 52°C. After incubation, the chip was taken out for the examination using a ZEISS Axio Observer fluorescence microscopy connected with ZEISS Axio Cam 305 and X-Cite 120Q fluorescence lamp illumination. For each chip’s microscopy, the same parameters were set up including 5× magnification objective, 10× magnification eyepiece, 700 ms exposure time, 2.1 gamma value, and 2000 white value. Six distinct regions without overlapping areas were randomly captured by the microscopy to cover about 2809 microreactions. The number of positive spots was counted by using the ImageJ software. The step-by-step operation is described below: Image>Type (8-bit)>Edit>Invert>Image>Adjust (Threshold: 0 and 245)>Apply>Analyze>Analyze Particles>Distribution>List. In the list, the count for over 0.02 bin start was enrolled and summed up.

Similarly, specificity assay of the digital WS-CRISPR assay was performed by testing the control plasmids mentioned above and sensitivity was evaluated by testing serially diluted SARS-CoV-2 RNA (Twist Bioscience) in water with concentrations of 5×10^5^, 5×10^4^, 5×10^3^, 5×10^2^, 5×10^1^ and 5×10^0^ copies/μl, as well as 3×10^6^ copies/μl SARS-CoV-2 RNA extracted from saliva sample. SARS-CoV-2 in each clinical sample was quantified using the calibration curve of digital WS-CRISPR by applying the averaged percentage of positive spots in six micrographs. Direct SARS-CoV-2 detection in saliva samples by digital WS-CRISPR assays were assessed through testing five saliva mock samples. The reaction system and procedure were the same as described above, just replacing the target RNA solution with 1.5 μl of heat-treated saliva sample solution at 95 °C for 5 min. The saliva mock samples (400 μl) were prepared by adding 1× inactivation reagent (0.0115 N NaOH, 1.0 mM EDTA, and 1.0 mM TCEP-HCl), 360 μl human saliva, and various volume percentages (0%, 1%, 2.5%, 5%, and 10%) of heat-inactivated SARS-CoV-2.

### Real-time quantitative RT-PCR assay

Real-time quantitative RT-PCR (RT-qCPR) assay for SARS-CoV-2 detection was carried out according to U.S. CDC 2019-Novel Coronavirus (2019-nCoV) Real-Time RT-PCR Diagnostic Panel (https://www.fda.gov/media/134922/download) with minor modifications. A typical RT-qPCR reaction system included 2 U/μl SuperScript IV reverse transcriptase, 1× QuantStudio master mix (Catalog # A26358), 0.5 μM nCOV_N1 forward primer, 0.5 μM nCOV_N1 reverse primer, 0.125 μM nCOV_N1 probe, and 1.5 μl of the target solution. The thermal cycler protocol consisted of Stage 1 (2.0 min at 25°C), Stage 2 (15.0 min at 50°C), Stage 3 (2.0 min at 95°C) and Stage 4 (40 cycles of 3.0 s at 95°C and 30 s at 55°C). The capture point of fluorescence was set at 55°C in Stage 4. Real-time quantitative analysis was performed in the Bio-Rad CFX96 Touch Real-Time PCR Detection System (Hercules, CA). Through testing serially diluted SARS-CoV-2 RNA (Twist Bioscience) with concentrations of 5×10^5^, 5×10^4^, 5×10^3^, and 5×10^2^ copies/μl, a four-point calibration curve was plotted based on the relationship between Cq value and the log of target concentration (**Supplementary Fig. 6**). On the basis of this curve, SARS-CoV-2 in each clinical sample was quantified by loading the determined Cq value.

### Statistics and reproducibility

GraphPad Software Prism 8.0.1 was used to plot real-time fluorescence curves, analyze linear regression, and verify statistical significance between two assay groups. The unpaired two-tailed *t*-test was made with the *p* value < 0.05 as the threshold for defining significance. For endpoint imaging of the chip using fluorescence microscopy, six distinct regions without overlapping areas were randomly taken to cover about 2809 microreactions. Unless otherwise specified, each image for visual detection or micrograph for chip testing is a representative of at least three independent experiments. To plot the linear relationship between percentage of positive spots and concentration of targets in digital WS-CRISPR, total positive spots in all the six micrographs were used and three chips were taken to run three independent assays. Clinical sample testing by both digital WS-CRISPR and RT-qPCR assays was repeatedly conducted at three times to ensure the data accuracy.

## Data Availability

All data needed to evaluate the conclusions in the paper are present in the paper and/or the Supplementary information. Additional data related to this paper may be requested from the authors.

## Acknowledgements

This research was supported, in part, by NIH R01EB023607, R61AI154642, and R01CA214072.

## Author information

### Contributions

X.D. and C.L. conceived the technique, performed experiments, analyzed the data, and drafted the paper. K.Y. and Z.L. contributed to data collection and data review. M.M.S. contributed to clinical samples, scientific advice, and resources for this study, and editing of the paper. C.L. supervised the whole project. All authors reviewed and approved the paper.

### Corresponding author

Correspondence to Dr. Changchun Liu

## Ethics declarations

X.D. and C.L. are inventors of a patent application filed by University of Connecticut based on this work.

## Supplementary information

Supplementary information for this article is available at the website.

## Supplementary information

**Supplementary Fig. 1.**
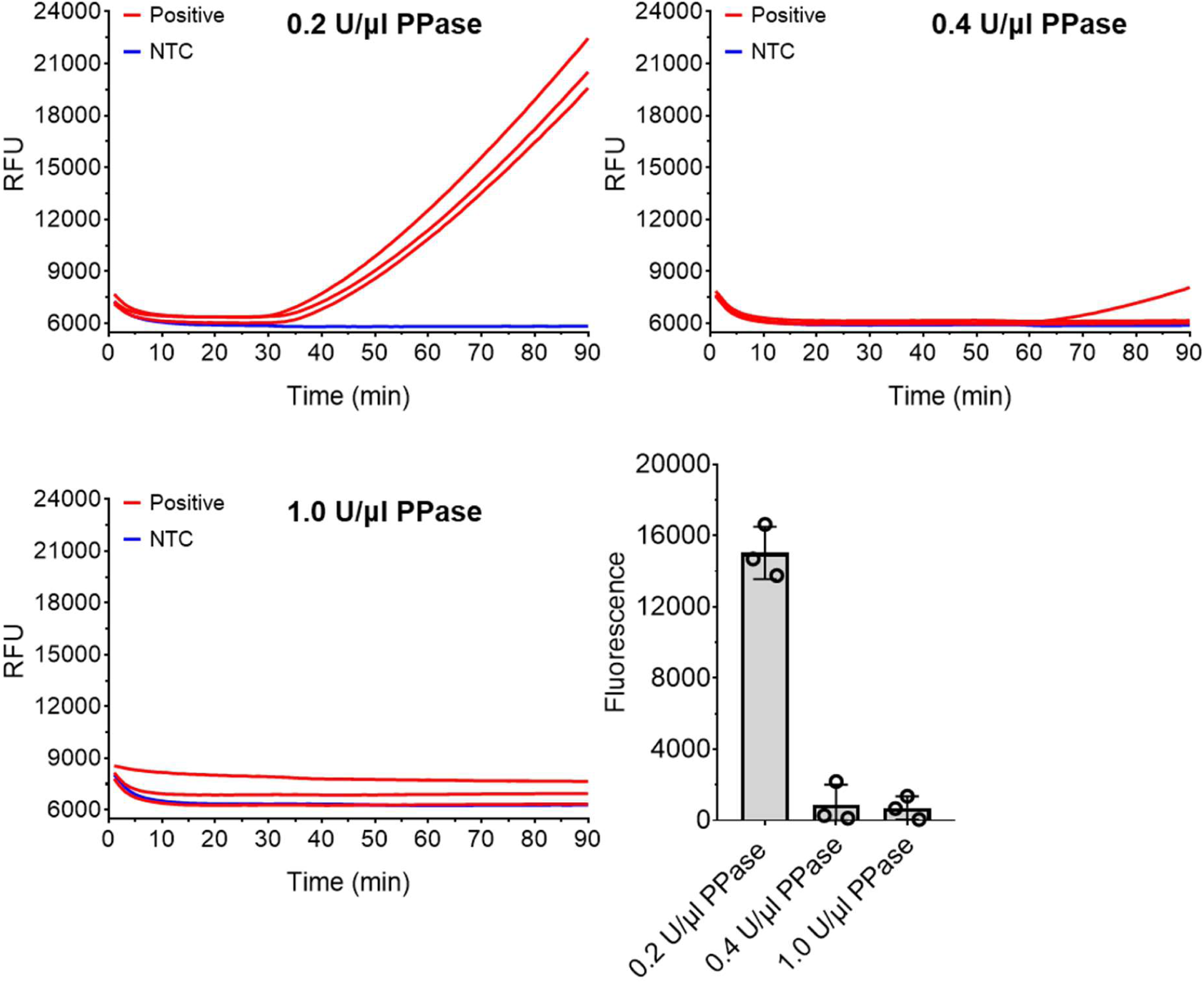
Effect of pyrophosphatase (PPase) concentration on one-pot WS-CRISPR assay. Positive, the reaction with 5×10^4^ copies/μl SARS-CoV-2 RNA. NTC, non-template control. Three replicates were run (n = 3). Error bars represent the means ± standard deviation (s.d.) from replicates.

**Supplementary Fig. 2.**
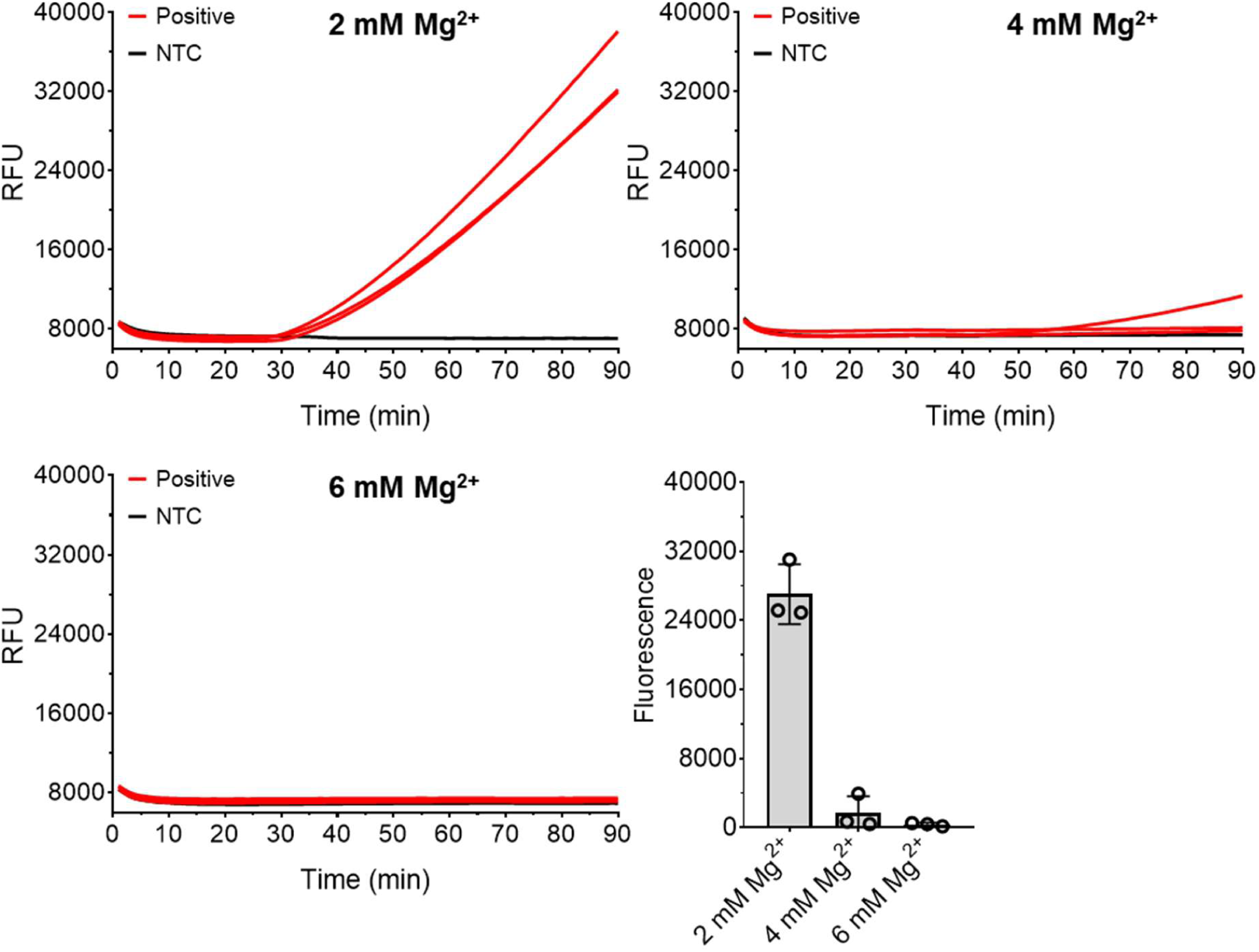
Effect of magnesium ion (Mg^2+^) concentration on one-pot WS-CRISPR assay. Positive, the reaction with 5×10^5^ copies/μl SARS-CoV-2 RNA. NTC, non-template control. Three replicates were run (n = 3). Error bars represent the means ± standard deviation (s.d.) from replicates.

**Supplementary Fig. 3.**
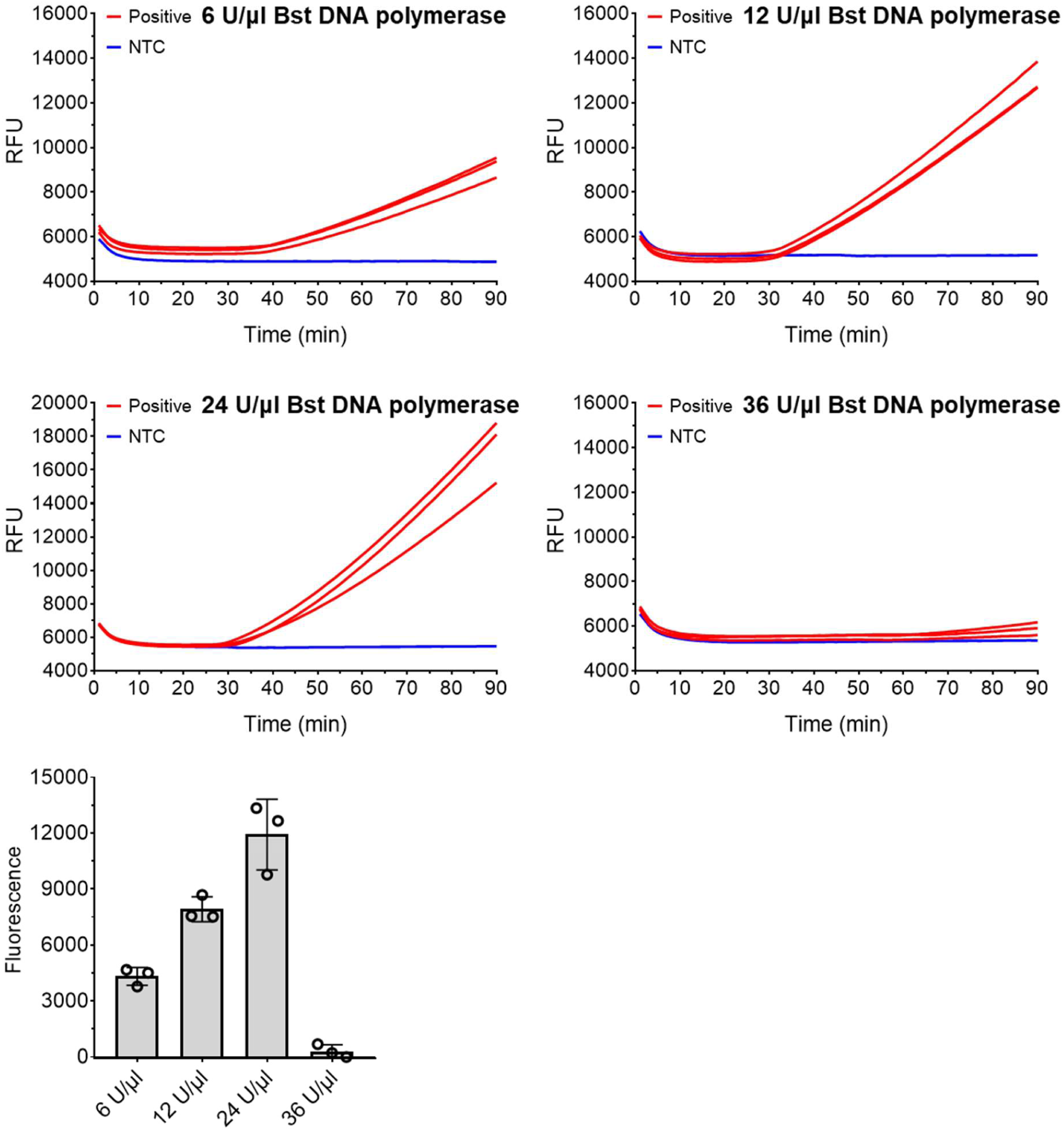
Effect of *Bst* DNA polymerase (large fragment) concentration on one-pot WS-CRISPR assay. Positive, the reaction with 5×10^4^ copies/μl SARS-CoV-2 RNA. NTC, non-template control. Three replicates were run (n = 3). Error bars represent the means ± standard deviation (s.d.) from replicates.

**Supplementary Fig. 4.**
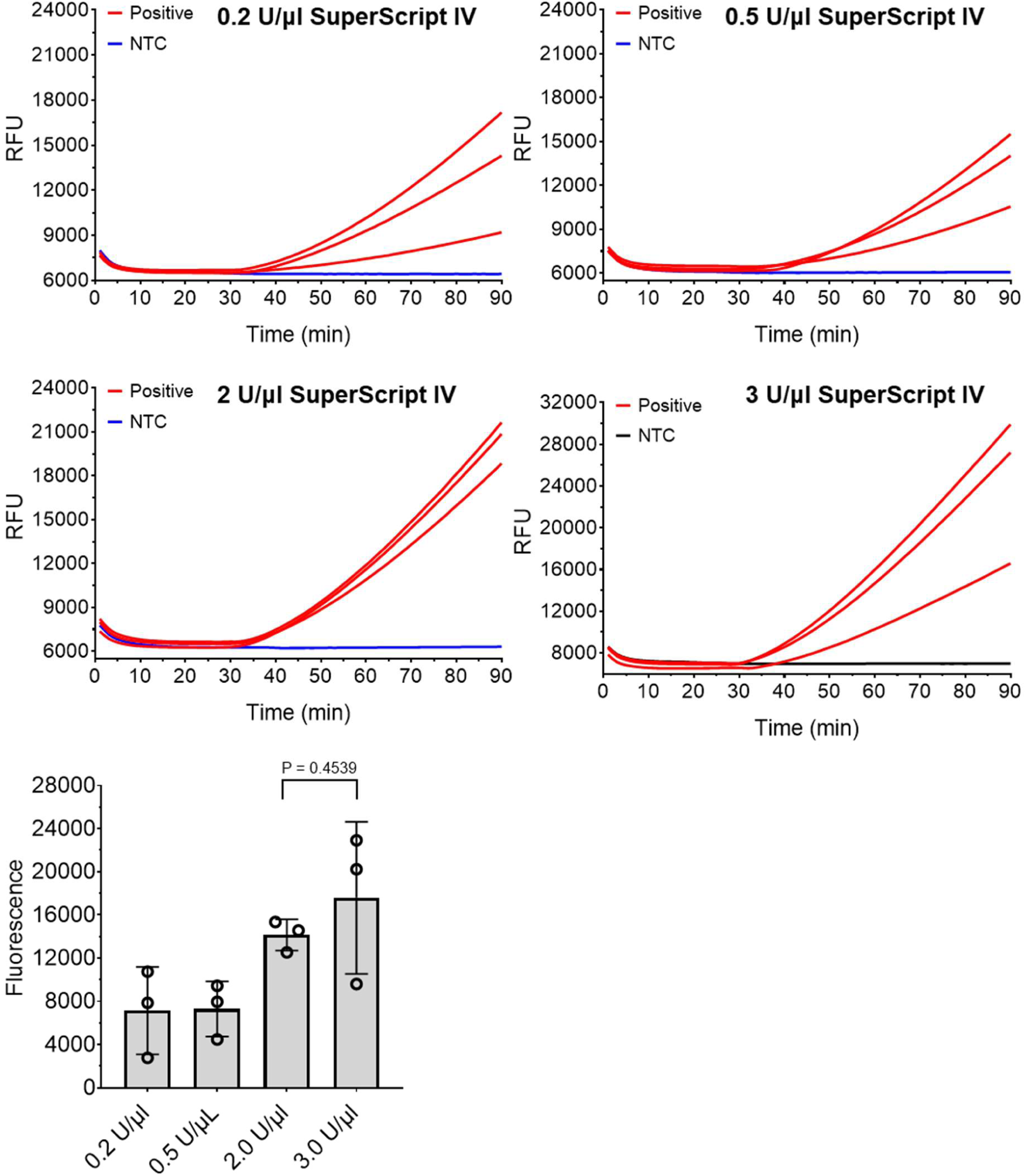
Effect of SuperScript IV reverse transcriptase concentration on one-pot WS-CRISPR assay. Positive, the reaction with 5×10^4^ copies/μl SARS-CoV-2 RNA. NTC, non-template control. Three replicates were run (n = 3). Error bars represent the means ± standard deviation (s.d.) from replicates. The statistical significance was analyzed using unpaired two-tailed t-test.

**Supplementary Fig. 5.**
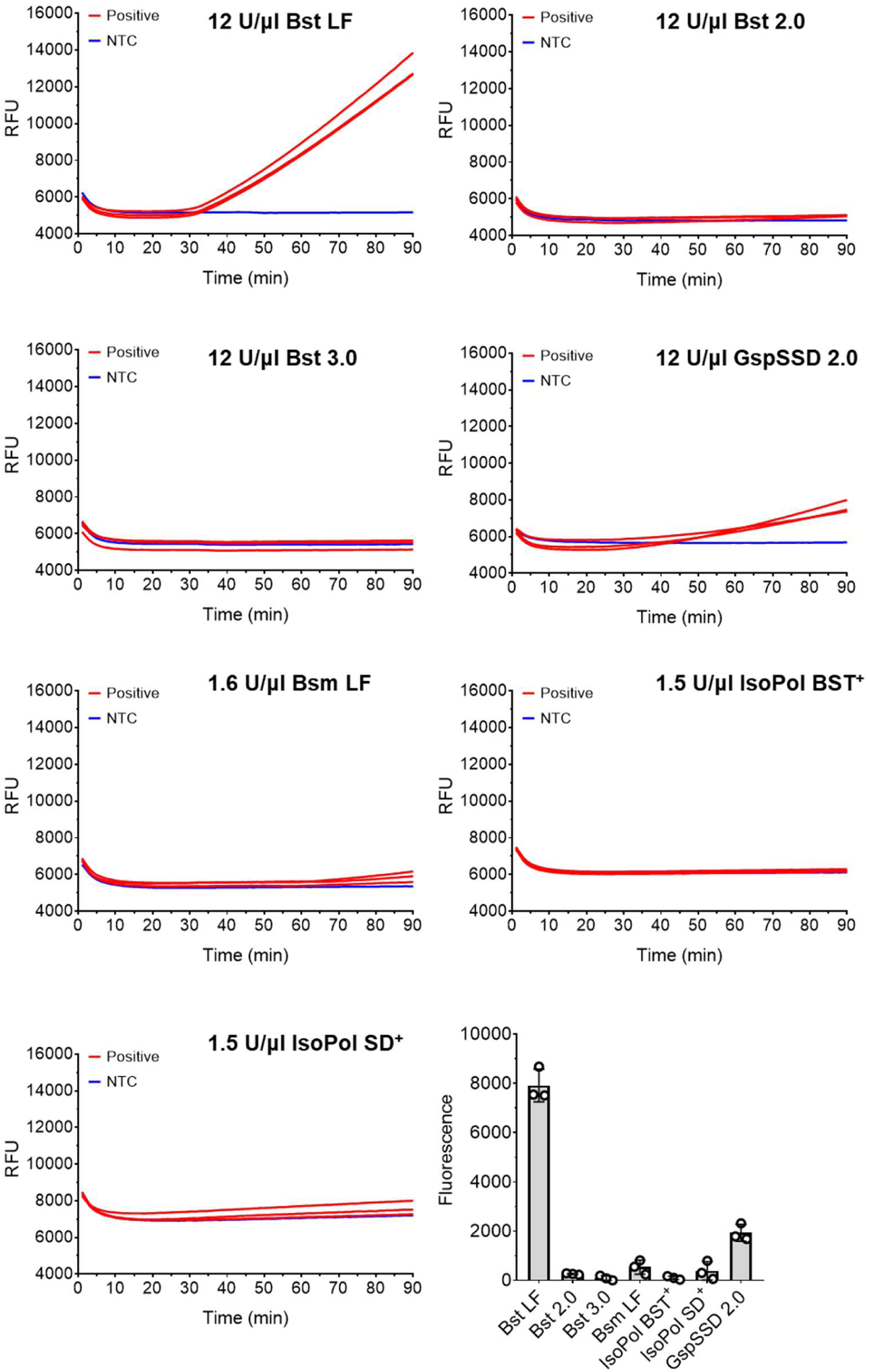
Effect of different DNA polymerases on one-pot WS-CRISPR assay. Positive, the reaction with 5×10^4^ copies/μl SARS-CoV-2 RNA. *Bst* LF, *Bst* DNA polymerase (large fragment). *Bst* 2.0, *Bst* 2.0 DNA polymerase. *Bst* 3.0, *Bst* 3.0 DNA polymerase. GspSSD 2.0, GspSSD 2.0 DNA polymerase. *Bsm* LF, *Bsm* DNA polymerase (large fragment). IsoPol BST^+^, IsoPol BST^+^ DNA polymerase. IsoPol SD^+^, IsoPol SD^+^ DNA polymerase. NTC, non-template control. Three replicates were run (n = 3). Error bars represent the means ± standard deviation (s.d.) from replicates.

**Supplementary Fig. 6.**
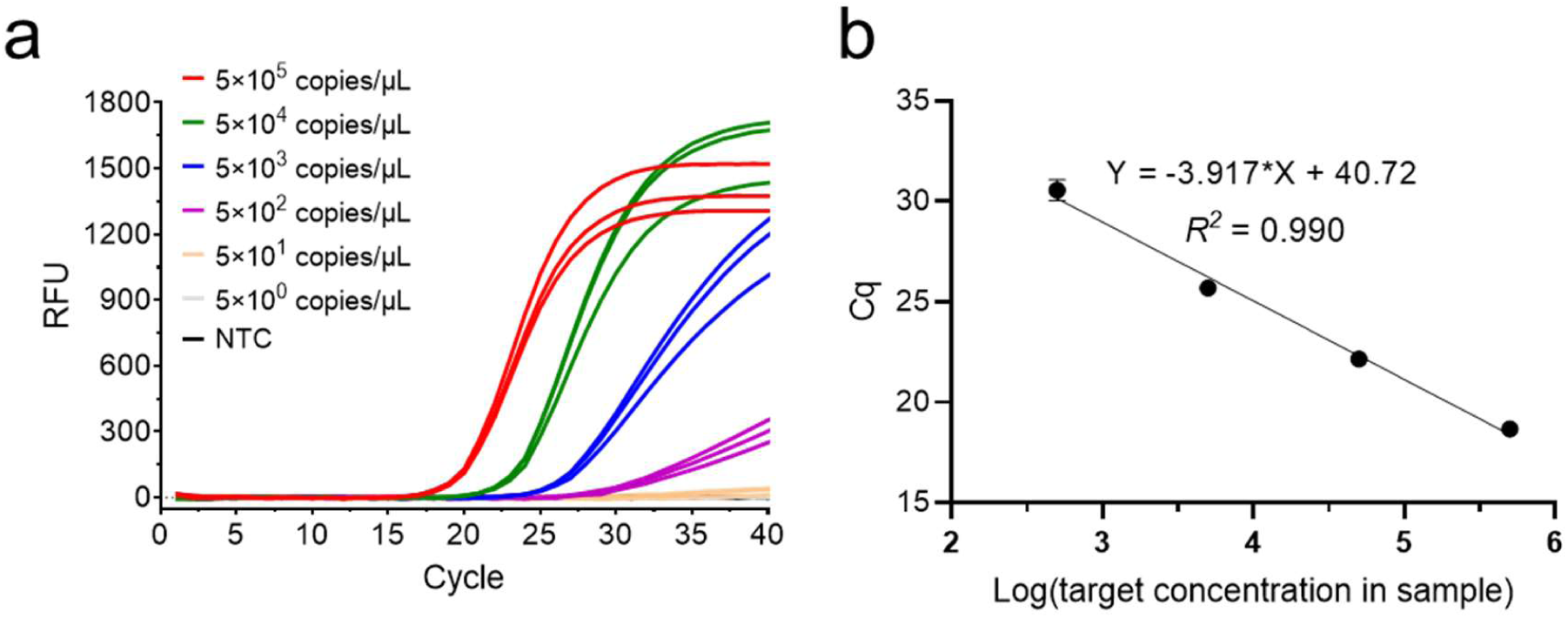
Real-time RT-qPCR assay of the tenfold serial dilution of SARS-CoV-2 RNA. **a**. Real-time fluorescence curves of the RT-qPCR assay. **b.** A four-point calibration curve developed to quantify the amount of SARS-CoV-2 RNA extracted from clinical samples. NTC, non-template control. Three replicates were run (n = 3). Error bars represent the means ± standard deviation (s.d.) from replicates.

**Supplementary Table 1.**
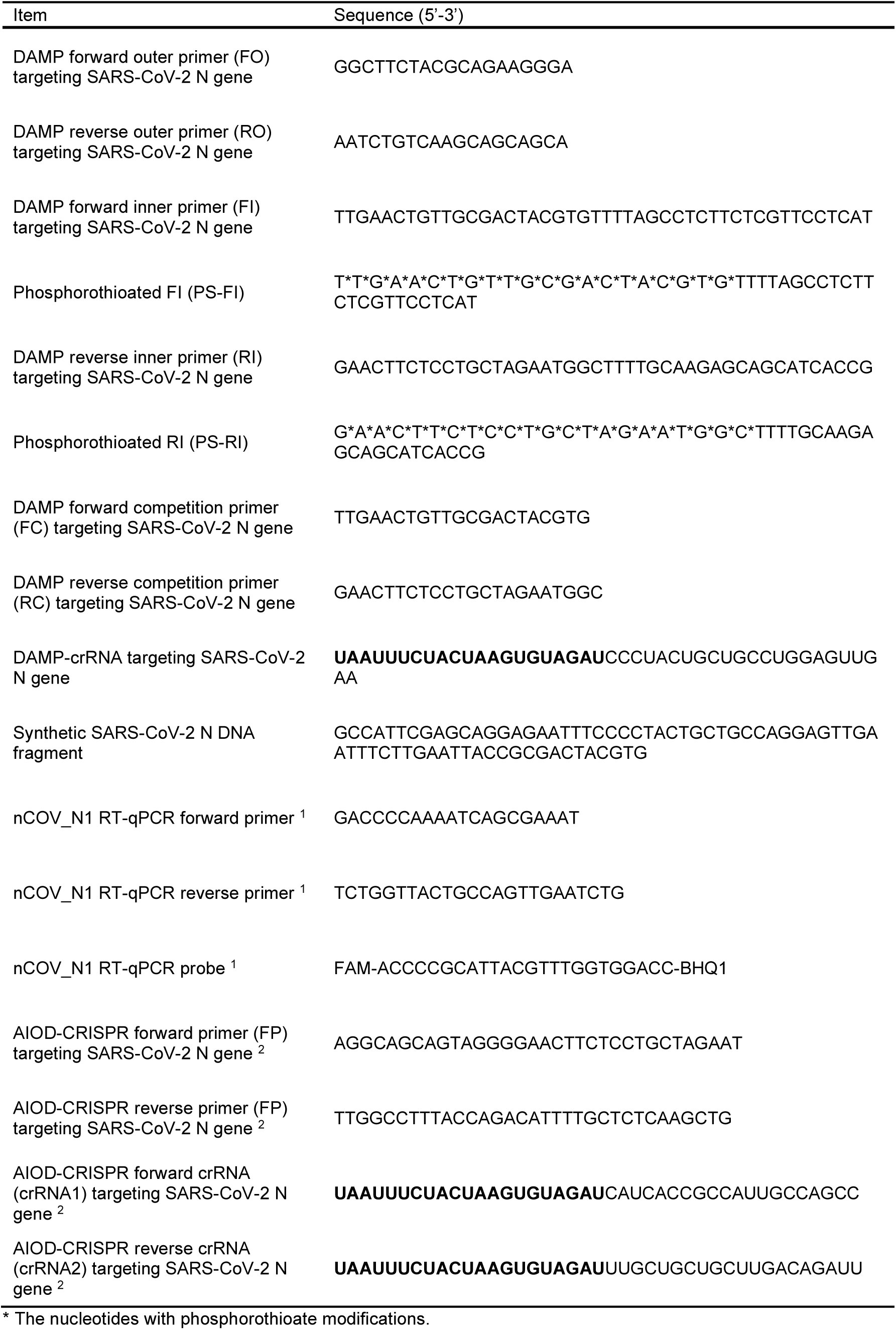
The list of all used sequences in this study

## Notes

### Competing Interest Statement

The authors have declared no competing interest.

### Author Declarations

The clinical samples without heat treatment and RNA extraction to inactivate the viruses were not involvoed. All clinical samples were de-identified RNA extracts and handled in compliance with ethical regulations and the approval of Institutional Review Board of the University of Connecticut Health Center (protocol #: P61067).

